# Trends in Prevention of Mother-to-Child Transmission of HIV at PEPFAR-Supported Sites in Mozambique, 2017–2023

**DOI:** 10.1101/2024.11.20.24317608

**Authors:** Judite Langa, Maria Inês de Deus, Sarah Arciniegas, Argentina Wate, Erica Bila, Arla Alfândega, Nelice Mate, Irene Rungo, Gizela Azambuja, Kwalila Tibana, Námita Eliseu

## Abstract

**Background:** Mozambique has implemented Prevention of Vertical Transmission (PVT) of HIV as part of national programming since 2002. Despite gains, vertical transmission rates remain high, estimated at 9% in 2023. We describe PVT progress at PEPFAR-supported sites.

**Methods:** We analyzed routine data from 656 PEPFAR-supported sites reported for fiscal years (October–September) 2017–2023. We calculated the proportion of pregnant women (PW) at first antenatal care (ANC1) who knew their status, HIV test positivity (positive tests divided by the total number of tests conducted); the proportion of PW on antiretroviral therapy (ART); the proportion of PW on ART with documented viral load (VL) test; the proportion of PW and breastfeeding women (BFW) with viral load suppression (VLS, defined as <1,000 copies/mL); the proportion of HIV-exposed infants (HEI) tested by 12 months who tested HIV positive; and the proportion of HEI-positives on ART. Data were analyzed by fiscal year.

**Results:** During 2017–2023, the proportion of PW who knew their HIV status at ANC1 increased from 90.4% (863,181/954,933) to 99.9% (1,047,870/1,049,225); HIV-positivity among PW at ANC1 decreased from 9.7% (84,038/863,181) to 8.1% (84,429/1,047,870); PW on ART remained above 99%; the proportion of PW who received a VL test increased from 24.5% (9,862/40,293) to 74.9% (46,292/61,818); and VLS increased from 51.4% (5,073/9,862) to 88.5% (40,950/46,292) and 60.8% (8,643/14,205) to 92.7% (149,893/161,695) among PW and BFW, respectively. During 2017–2023, HEI testing coverage increased from 76.5% (64,322/84,038) to 96.0% (82,689/86,132), and HEI HIV positivity declined from 8.5% (5,487/64,322) to 2.5% (2,075/82,689); during 2018–2023, the proportion of HIV positive HEI on ART increased from 82.7% (3,521/4,256) to 93.3% (1,936/2,075).

**Conclusion:** PEPFAR-supported sites have achieved progress in PVT. Continued implementation of sound interventions to improve ART initiation and retention, VLS, and HEI treatment linkage can lead to further reductions in vertical transmission and the elimination of mother-to-child transmission of HIV as a public health problem in the country.

## Introduction

Vertical transmission (VT) of HIV continues to present a severe challenge to epidemic control. In 2010, WHO estimated that more than 90% of new HIV infections among children occur due to MTCT.^1^ While VT is preventable without intervention, the estimated risk of transmission during pregnancy, birth, and breastfeeding are 5–10%, 10–20%, and 5–20%, respectively.^2^ The Joint United Nations Programme on HIV and AIDS (UNAIDS) 2030 goals and the World Health Organization (WHO) Triple Elimination Initiative set a VT rate benchmark of <5% in breastfeeding populations.^3^

In 2002, Mozambique began the prevention of vertical transmission (PVT) program, using single-dose nevirapine for prophylaxis in selected sites. The program continuously expanded and, in 2013, implemented the Option B+ Strategy, through which all pregnant and breastfeeding women (PBFW) were provided with lifelong antiretroviral therapy (ART) upon diagnosis, irrespective of HIV clinical stage or CD4 cell count.^4^ In 2016, dolutegravir (DTG) based regimens were introduced as part of Option B+ programming, as they achieve more rapid VLS and are more effective in achieving VLS during pregnancy^5^ In 2019, enhanced prophylaxis with nevirapine and zidovudine syrups for the exposed infant were introduced. As the program developed, PVT data capture has also evolved. For example, longitudinal antenatal care (ANC) and at-risk child consultation (ARCC) registers were rolled out in 2015 and 2016. During the same period, the program reinforced interventions to improve maternal adherence and retention in care, which included the introduction of peer support through mentor mothers (MM), who are women living with HIV who went through the PVT program and are willing to help others in the same situation. As of August 2023, there were 3,805 MM deployed in 96% of sites supported by U.S. President’s Emergency Plan for AIDS Relief (PEPFAR),^6^ working to improve HIV outcomes for HIV-positive mothers and their exposed infants.

The Elimination of Vertical Transmission (EVT) of HIV is one of the core strategies of Mozambique’s Plan for Triple EVT of HIV, Syphilis and Hepatitis 2020-2024, which aims to reduce HIV transmission from mother to child to <5% and the incidence of new pediatric infections to <750 per 1,000 live births. The EVT program focuses on four core interventions: 1) biomedical interventions, including HIV testing and linkage to ART; 2) promotion of responsible paternity through interventions to strengthen male engagement in maternal and child health care, improve viral load suppression (VLS) among PBFW and utilization of viral load (VL) results to inform gestation planning; 3) sustaining VLS among women of reproductive age, including PBFW, through psychosocial support and 4) early infant HIV diagnosis, antiretroviral (ARV) prophylaxis for HIV-exposed infants (HEI) and linkage to ART for HIV-positive infants.^7^

In 2004, PEPFAR began supporting Mozambique, including PVT programming. In this article, we describe PVT progress at PEPFAR-supported sites during 2017–2023, focused on describing antenatal care (ANC) coverage in the general women’s population; the PVT cascade among PBFW; and the diagnostic and treatment cascade among HEI.

## Methods

We used PVT data from the Monitoring, Evaluation, and Reporting (MER) database, which contains data reported from PEPFAR-supported sites, for 2017-2023. Some programmatic data was not available from 2017, so periods for those indicators started based on data availability. Additionally, UNAIDS Spectrum modeling^8^ and the National Demographic and Health Data Survey (DHS)^9^ were used for national estimates on ANC visits and VT rates. Within our analysis period, the DHS only reported on indicators for 2022-2023. Spectrum was also utilized to report data from 2023.

Among PBFW, we analyzed: the estimated proportion of pregnant women (PW) attending ANC; the proportion of PW attending first ANC visit (ANC1) who knew their HIV status, disaggregated by known HIV-positive status at entry and newly tested HIV positive in ANC1; the proportion of PW who attended ≥4 ANC visits (ANC4); the proportion PBFW retested for HIV after ANC1, calculated as the number of PBFW who were HIV-negative at ANC1 and were retested post-ANC1 over the number of PW who were HIV-negative at ANC1; the proportion of HIV-positive PW at ANC1 on ART; the number of VL tests among PBFW on ART; VL coverage, or the proportion of pregnant women documented as “Already on ART” in the past 12 months with a VL test; and the proportion of PBFW on ART with VLS (defined as viral load test results <1,000 copies/mL). National PVT efforts are especially focused on improving outcomes among PBFW ages 10-24 years old; however, programmatic, viral load outcomes among PBFW are not further disaggregated by age group when reported to DATIM. Because of this, we also analyzed VLS among the general population of female PLHIV aged 10-24 years old and 25+ on ART.

We used the total number of HIV-positive PBFW as a proxy measure for the number of HEI. Among HEI, we analyzed: HEI testing coverage, calculated as the number of HEI tested for HIV by age 12 months over the number of HIV-positive PBFW; positivity rate, calculated as the number of positive tests over the total number of tests conducted among HEI; VT rate, calculated as the number of HIV-positive HEI over the number of mothers in PVT; linkage to treatment rate, calculated as the number of HIV positive HEI initiated on ART over the number of positive tests; and HEI with final outcomes, defined as those who had a documented final outcome (i.e., HIV-infected, HIV-uninfected, HIV-final status unknown, or died without status known) by age 18 months.

Descriptive statistics were used to analyze annual trends, with the start date depending on data availability. Data were analyzed by fiscal year (October-September), province, and age groups (PBFW and women on ART: 10-24 years, ≥25 years; HEI: <2 months, 2-12 months). Age disaggregations for PBFW and women on ART were based on definitions for who are considered youths and adolescents, a mentioned target programmatic group. Additionally, age categories of < 2 months versus 2-12 months were used because of national PVT guidelines, recommending testing for HEI in the first two months of infancy. Viral load data were analyzed by pregnancy and breastfeeding status; HEI final outcomes data were disaggregated by outcome type. Data were analyzed using PowerBI software.

This activity was reviewed by CDC, deemed not research, and was conducted consistent with applicable federal law and CDC policy.^1^

## Results

### Pregnant and Breastfeeding Women

According to national DHS estimates, among PW aged 15-49 years old in 2022-2023, 87.3% attended ANC1 and 48.6% attended ≥4 ANC visits.^9^ ANC coverage was higher in the southern provinces, with the highest ANC coverage in Maputo Province (ANC1: 99.5%; ANC4: 82.7%) and Maputo City (ANC1: 100.0%; ANC4: 81.2%). The remaining results on PBFW come from routinely collected programmatic data. The proportion of PW who knew their HIV status at ANC1 increased from 90.4% (863,181/954,933) in 2017 to 99.9% (1,047,870/1,049,225) in 2023. The proportion of PBFW who were HIV-negative at ANC1 and were retested for HIV post-ANC1 increased from 22.1% (42,374/192,202) to 83.7% (201,938/241,328) during June-September 2019 and June-September 2023, respectively. Between 2017 and 2023, the proportion of HIV-positive women in ANC1 decreased from 9.7% (84,038/863,181) to 8.1% (84,429/1,047,870). HIV-positivity decreased across all age groups, especially among 10–14-year-olds (9.1% to 2.0%) and 20–24-year-olds (12.9% to 5.8%). Maputo Province had the largest decrease in positivity among PW (21.4% to 9.7%). Among the HIV-positive PW in ANC1, the proportion who were newly identified as HIV-positive (as opposed to known status at entry) decreased from 52.3% (43,937/84,038) to 26.8% (22,621/84,429) (Figure 1).

**Figure 1.**
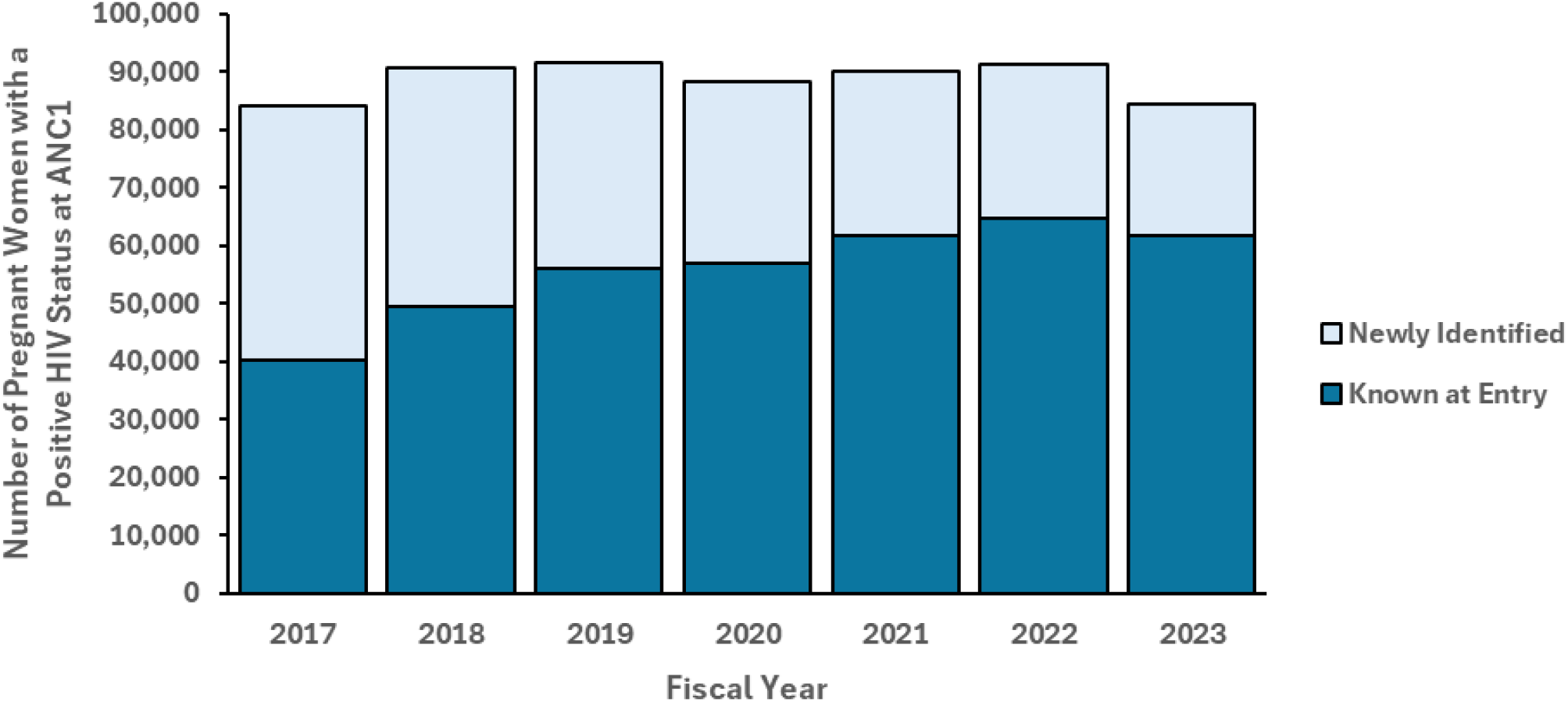
Trends Among HIV-Positive Women Attending ANC1 at PEPFAR-Supported Sites, Newly Identified vs. HIV Status Known at Entry, Mozambique, 2017-2023

Since 2017, reported linkage to ART among HIV-positive women in ANC has been greater than 99%. From 2017 to 2023, the number of VL tests conducted among PBFW increased 7.6 times, from 24,067 to 207,987. The proportion of pregnant women who received a VL test increased from 24.5% (9,862/40,293) to 74.9% (46,292/61,818) during June-September 2017 and June-September 2023, respectively. For this same period, the VLS rate increased from 60.8% (8,643/14,205) to 92.7% (149,893/161,695) among BFW and from 51.4% (5,073/9,862) to 88.5% (40,950/46,292) among PW, with VLS rates consistently higher among breastfeeding women (Figure 2). Among female PLHIV aged 10–24 years on ART, the VLS rate increased from 57.5% (10,445/18,159) to 88.5% (98,049/110,805), and VLS suppression among female PLHIV aged ≥25 years increased from 69.4% (44,773/64,556) to 94.7% (724,080/764,550).

**Figure 2.**
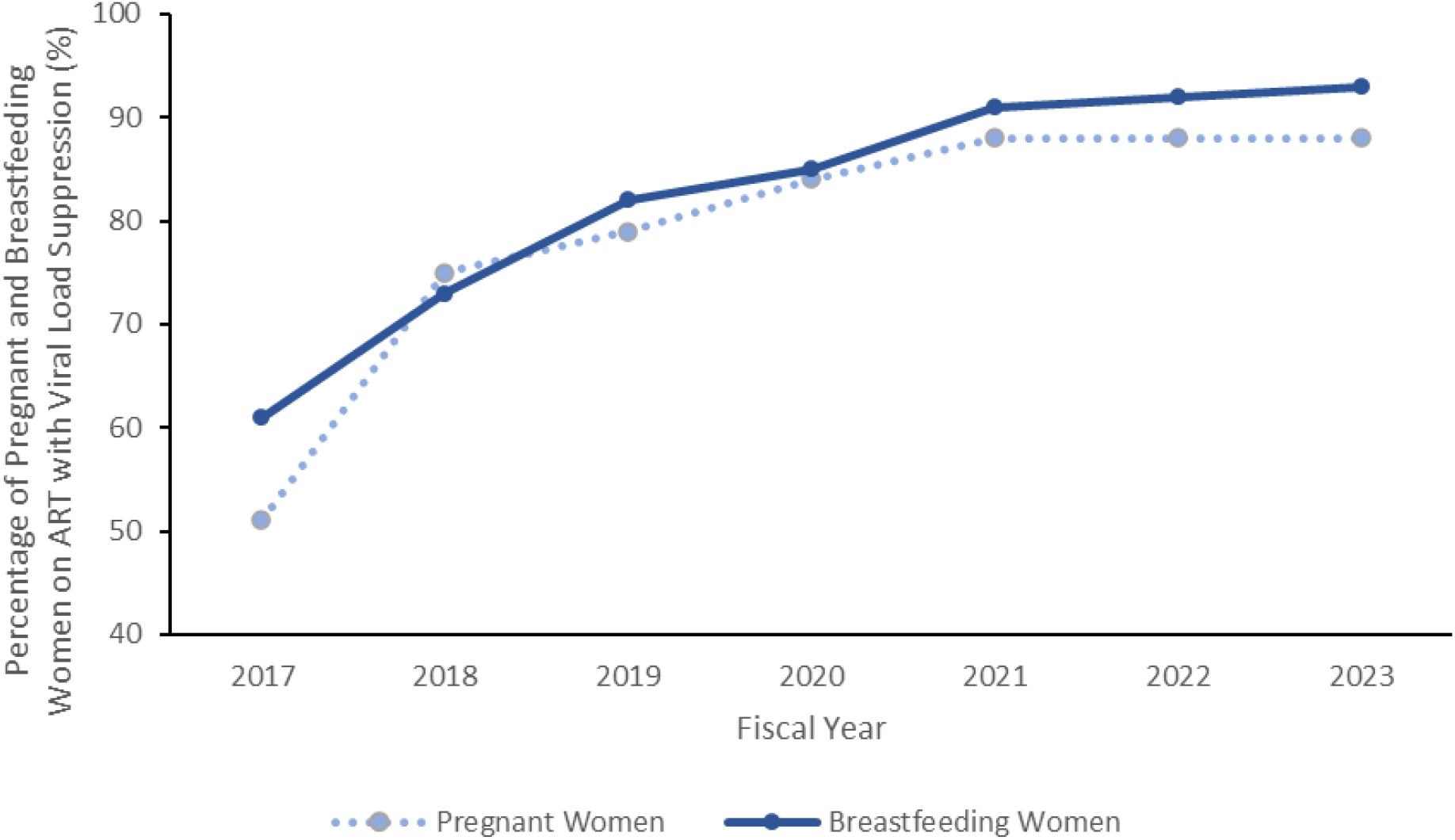
Viral Load Suppression Rate Percentages Among Female PLHIV on ART in ANC at PEPFAR-Supported Sites, Mozambique, 2017-2023

### HIV Exposed Infants

Between 2017 and 2023, the estimated number of HEI increased from 84,038 to 86,132. During this period, HEI testing coverage increased from 76.5% (64,322/84,038) to 96.0% (82,689/86,132). Over time, the proportion of HEI receiving their first HIV test between at <2 months of age increased, with 70.4% (45,252/64,322) of infants tested falling within this age category in 2017 compared to 93.3% (77,182/82,689) in 2023. In addition, the test positivity rate among HEI decreased from 8.5% (5,487/64,322) to 2.5% (2,075/82,689). All provinces had a decreased test positivity rate except Tete, which increased from 1.0% (19/1,929) to 2.4% (91/3,851). In 2023, the lowest HEI test positivity rate was in the three southernmost provinces (Maputo City: 1.5% (72/4,792); Maputo Province: 1.3% (83/6,555); Gaza: 1.7% (114/6,673). Between 2018 and 2023, HEI test positivity rate decreased for <2 -month-olds (2018: 3.8% [2,337/61,113]; 2023: 1.5% [1,158/77,182]), while positivity increased among 2–12-month-olds (2018: 13.7% [1,919/13,966]; 2023: 16.7% [917/5,507). Among HEI diagnosed with HIV, the percent linkage to treatment increased from 82.7% (3,521/4,256) in 2018 to 93.3% (1,936/2,075) in 2023, with increases among both those aged <2 -months (83.8% [1,958/2,337]; 93.8% [1,086/1,158]) and those aged 2–12 months (81.4% [1,563/1,919]; 92.7% [850/917]). Among HEI with registered final outcomes in 2019 and 2023, there was a reduction in the proportion of final outcomes registered as HIV-final status unknown (23.9% [14,724/61,705] to 8.9% [6,879/ 77,151]), HIV-infected (5.7% [3,535/61,705] to 2.8% [2,136/77,151]), or having died (0.9% [564/61,705] to 0.7% [539/77,151]) (Figure 3). According to Spectrum estimates, the MTCT rate decreased nationally from 13.3% in 2017 to 8.7% in 2023.^8^

**Figure 3.**
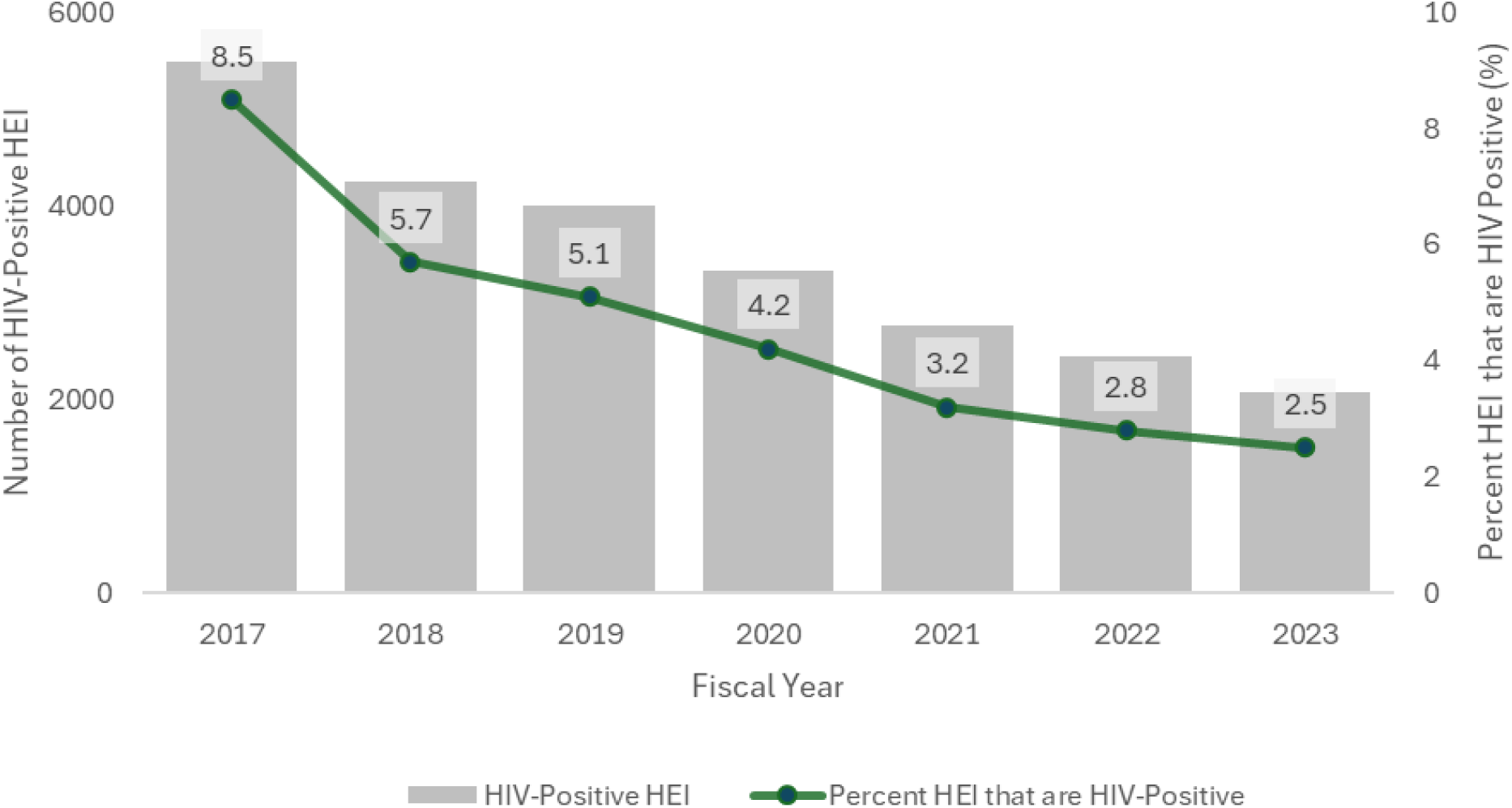
Trends in HIV Positivity Among HEI at PEPFAR-Supported Sites, Mozambique, 2017— 2023

## Discussion

Since 2017, there has been tremendous success in PVT at PEPFAR-supported sites, with linkage to ART among PBFW remaining above 99%, VLS rate among PW reaching 88%, and test positivity rate among HEI decreasing to 2.5%. These improvements might have contributed to better outcomes for both mothers and babies, including the prevention of new infections among children, who would otherwise require life-saving ART.^3^

Despite high HIV testing coverage among PW and linkage to ART among PBFW, there were still missed opportunities for preventing vertical transmission, and the country has lagged in reaching the full set of indicator benchmarks outlined in the bronze tier of the WHO’s Path to Elimination (specifically, 90% coverage in ANC1, HIV testing, and ART linkage). ^10^ Indeed, by the end of the analysis period, ANC1 coverage was still lower than 90%, nationally, with significantly lower coverage of 4 or more visits.^9^ ANC visits are essential for the detection of HIV among pregnant women; therefore, challenges in ANC attendance are a significant barrier for PVT. In terms of VLS, there were greater challenges in achieving suppression among pregnant women on ART compared to breastfeeding women on treatment, who reached 88% and 93% VLS by 2023, respectively. These results are aligned with findings from other African countries, which have shown a similar pattern of higher VLS among BFW.^11-13^ Our results show that an increasing proportion of PW living with HIV seen at first ANC attended this consultation already knowing their HIV-positive status (48% in 2017;73% in 2023), indicating improved overall HIV case finding. Nonetheless, routine programmatic data (not shown) has indicated that ART interruption often occurs among women before pregnancy and, added to general late ANC1 attendance (only 21% of PW arrive at gestational age <12 weeks^14^), may hinder timely VLS during this period and may be linked to socio-cultural as well as structural health system related factors. While improvements in VLS rate among female PLHIV aged 10-24 are encouraging, VLS has remained lower among this age group compared to female PLHIV ≥25 years, reaching 89% among those aged 10-24 years in 2023. The consistently lower VLS rate observed among female PLHIV aged 10-24 likely translates to poorer suppression among PBFW within the same age group. Additionally, studies have found poorer ART retention and care-seeking behaviors among younger mothers in Mozambique, which would be consistent with lower VLS among this age group.^15^ Opportunities for further improvements include strengthening PMTCT strategies, including HIV maternal retesting and use of HIV pre-exposure prophylaxis (PrEP) for PBFW at risk of HIV infection.

Additionally, there have been remaining gaps related to client behavior and knowledge, such as unknown HIV status prior to conception, unplanned pregnancy, late attendance to ANC, illiteracy and non-disclosure of HIV-positive status to partners.^13,16-18^ Health care providers may face challenges when there is insufficient training to provide quality ANC.^16^ Additionally, the volume of nurses or midwives in Mozambique is likely inadequate for effective ANC. While an estimated 83.5% of pregnant women in ANC were attended by a nurse or midwife in 2022-2023,^9^ the ratio of midwifery and nursing personnel to population was an estimated 5.12 per 10,000 people in 2022.^19^Lastly, the health system must overcome poor infrastructure, low coverage of health facilities within the population, and stock outs of consumables, among other issues.^18^

HEI follow-up, including PCR testing, has improved considerably in the country. Despite improvements over time, HEI were still being tested later in infancy (after 2 months of age) by the end of 2023. Missed opportunities in early infant PCR testing lead to later HIV-diagnosis, which may explain the observed higher HIV-positivity seen among 2-12 month-old HEI (16.7%).

There are several limitations. First, data analyzed were only from PEPFAR-supported sites from 2017 to 2023, and women in ANC and post-natal care at other facilities may have had different outcomes. We included all sites that received PEPFAR funding at any point during the review period, though not all sites received PEPFAR funding for the entire duration. Additionally, VL coverage among breastfeeding women cannot be calculated using MER data, which limits our understanding of VL testing and suppression among this target population. To better understand these differences in outcomes, we need to further investigate any challenges that may be facing PBFW and how the data are being collected and reported.

## Conclusion

PEPFAR-supported sites have achieved progress in PVT. Continued implementation of sound interventions to improve ART initiation and retention, VLS, and HEI treatment linkage can lead to further reductions in vertical transmission and the elimination of mother-to-child transmission of HIV as a public health problem in the country.

## Data Availability

All data produced are available from PEPFAR Monitoring, Evaluation, and Reporting program data.

https://data.pepfar.gov/datasets#PDD

## Acknowledgements

We are grateful to site level staff and implementing partners for the tremendous work they do in supporting and maintaining national PVT programming at the local level. Additionally, we acknowledge the years of guidance and partnership from the Mozambican Ministry of Health (MISAU).

## Conflict of Interest

All authors declare no competing interests.

## Funding Acknowledgement

This manuscript has been supported by the President’s Emergency Plan for AIDS Relief (PEPFAR), through the Centers for Disease Control and Prevention (CDC). The findings and conclusions in this manuscript are those of the authors and do not necessarily represent the official position of the funding agencies.

## Footnotes

1 See e.g., 45 C.F.R. part 46.102(I)(2), 21 C.F.R. part 56; 42 U.S.C. 241(d); 5 U.S.C. 552a; 44 U.S.C. 3501 et seq.

## Notes

### Competing Interest Statement

The authors have declared no competing interest.

### Funding Statement

This manuscript has been supported by the President's Emergency Plan for AIDS Relief (PEPFAR) through the U.S. Centers for Disease Control and Prevention (CDC). The findings and conclusions in this manuscript are those of the author(s) and do not necessarily represent the official position of the funding agencies.

### Author Declarations

The study used PEPFAR Monitoring, Evaluation, and Reporting program.

